# Age and smoking predict antibody titres at 3 months after the second dose of the BNT162b2 COVID-19 vaccine

**DOI:** 10.1101/2021.08.06.21261590

**Authors:** Yushi Nomura, Michiru Sawahata, Yosikazu Nakamura, Momoko Kurihara, Ryousuke Koike, Otohiro Katsube, Koichi Hagiwara, Seiji Niho, Norihiro Masuda, Takaaki Tanaka, Kumiya Sugiyama

**Affiliations:** Department of Respiratory Medicine and Clinical Immunology, National Hospital Organization Utsunomiya National Hospital; Department of Pulmonary Medicine and Clinical Immunology, Dokkyo Medical University; Division of Pulmonary Medicine, Department of Medicine, Jichi Medical University; Department of Public Health, Jichi Medical University; Department of Surgery, National Hospital Organization Utsunomiya National Hospital; Department of Orthopaedic Surgery, National Hospital Organization Utsunomiya National Hospital; Department of Respiratory Medicine and Clinical Immunology, Dokkyo Medical University Saitama Medical Center

**Keywords:** SARS CoV-2, viral infection, clinical epidemiology

## Abstract

**Objective:** We aimed to determine antibody (Ab) titres 3 months after the second dose of the BNT162b2 coronavirus disease-2019 (COVID-19) vaccine and to explore clinical variables predicting these titres in Japan.

**Methods:** We enrolled 378 healthcare workers (255 women, 123 men) whose blood samples were collected 91±15 days after the second of two inoculations of the BNT162b2 COVID-19 mRNA vaccine (Pfizer/BioNTech) given 3 weeks apart. Medical histories and demographic characteristics were recorded using a structured self-reported questionnaire. The relationships between Ab titres and these factors were analysed.

**Results:** Median age (interquartile range [IQR]) of the participants was 44 (32-54) years. Median Ab titre (IQR) against the severe acute respiratory syndrome coronavirus 2 (SARS-CoV-2) spike antigen was 764 (423-1140) U/mL. Older participants had significantly lower Ab titres; median (IQR) Ab titres were 942 (675-1390) and 1095 (741-1613) U/mL in men and women in their 20s, respectively, but 490 (297-571) and 519 (285-761) U/mL in men and women in their 60s-70s, respectively. In the age-adjusted analysis, the only risk factors for lower Ab titres were male sex and smoking. However, the sex difference may have arisen from the sex difference in smoking rate. Moreover, Ab titres were significantly lower in current smokers than in ex-smokers.

**Conclusion:** The most important factors associated with low Ab titres were age and smoking habit. In particular, current smoking status caused lower Ab titres, and smoking cessation before vaccination may improve the individual efficacy of the BNT162b2 vaccine.

## INTRODUCTION

In Japan, the BNT162b2 vaccine (Pfizer/BioNTech) was selected as the first coronavirus disease-2019 (COVID-19) mRNA vaccine to be administered to healthcare professionals, starting in February 2021. Severe acute respiratory syndrome coronavirus 2 (SARS-CoV-2) has an enveloped, single, positive-stranded RNA genome that encodes four major viral structural proteins, namely nucleocapsid (N), spike (S), envelope (E) and membrane (M) proteins; the latter three proteins are found in its membrane. The spike protein guides viral entry into host cells [1] by binding to ACE2 (angiotensin-converting enzyme 2), the main virus receptor, which is widely expressed on epithelial cells and macrophages [1-3] and is thus an ideal target for mRNA vaccine development [3].

Because efficacy in clinical trials and effectiveness in the community depend on the proportions of SARS-CoV-2 variants spreading in a given area, immunogenicity has attracted increasing attention as an individual index for the efficacy of COVID-19 mRNA vaccines. Humoral immunity plays major roles in protecting against and surviving SARS-CoV-2 infection [4, 5], and neutralising antibodies (Abs) are correlated with protection against several viruses, including SARS-CoV-2 [6-9]. However, few studies have investigated real-world Ab titres following vaccination with BNT162b2; instead, focus has been placed on the Ab status shortly after vaccination, which can provide vital information for predicting long-term effectiveness [10-12]. A few studies have investigated the associations between Ab titres and individual demographic, medical, and lifestyle factors [13]; these studies focused on short-term Ab titre data and demonstrated that lower Ab titres were associated with older age [10-12], male sex [10], obesity [14, 15], smoking habit [15], drinking habit [10], hypertension [15], cancer [16, 17], use of immunosuppressive drugs [10] and a longer time elapsed after vaccine inoculation [10, 15].

Therefore, in our preliminary study [18], we reported the first medium-term data on Ab titres against the SARS-CoV-2 spike antigen produced in response to this mRNA vaccine in the Japanese population. Our results showed that the average peak titre of 2031.7±692.0 U/mL in 6 healthcare workers in their 50s and 60s had markedly decreased to 513.3±261.7 U/mL by 15 weeks after the second inoculation. Here, we aimed to determine Ab titres against the SARS-CoV-2 spike antigen 3 months after the second dose of the BNT162b2 vaccine in 378 healthcare workers and to explore the factors associated with these Ab titres across a comprehensive range of clinical and lifestyle characteristics in Japan.

## METHODS

### Population and study design

In this single-centre prospective observational study, we recruited healthcare workers whose blood samples were collected 91±15 days after the second of two BNT162b2 vaccine inoculations (Pfizer/BioNTech) administered 3 weeks apart in February–March 2021 in National Hospital Organization Utsunomiya National Hospital in Tochigi prefecture, Japan.

Initially, 381 participants were recruited, but we excluded 1 participant who received only the first dose of the BNT162b2 vaccine and 2 participants whose blood sampling confirmed the presence of Abs against SARS-CoV-2 (nucleoproteins) prior to vaccination. Finally, we enrolled 378 healthcare workers (255 women, 123 men), including the 6 from our preliminary study [18], and their medical histories and demographic characteristics were recorded using a structured self-reported questionnaire.

Blood samples collected 91±15 days after the second inoculation were used to measure total Ab titres against the SARS-CoV-2 spike antigen, using a commercially available electrochemiluminescence immunoassay (ECLIA) (Elecsys® Anti-SARS-CoV-2 RUO; Roche Diagnostics) [19]. The relationships between Ab titres against the SARS-CoV-2 spike antigen and clinical and lifestyle characteristics were analysed.

This study was approved by the Ethics Committee of National Hospital Organization Utsunomiya National Hospital (No. 03-01, April 19, 2021). Written informed consent was obtained from all study participants before enrolment.

### Data analysis

Nonparametric continuous data are expressed as the median with interquartile range (IQR). Categorical data are presented as absolute numbers and relative frequencies (*n*, %). To calculate Spearman’s rank correlation coefficient and perform the Mann-Whitney *U* test, we used Statistical Package for the Social Sciences (SPSS version 25).

## RESULTS

### Study population

In total, 378 healthcare workers (255 women, 123 men) were enrolled in this study. Their baseline characteristics are summarised in **Table 1**. Briefly, median age (IQR) of the participants was 44 (32-54) years. Nurses (n=177) and physicians (n=38) comprised 56.9% of the study population.

**Table 1.**
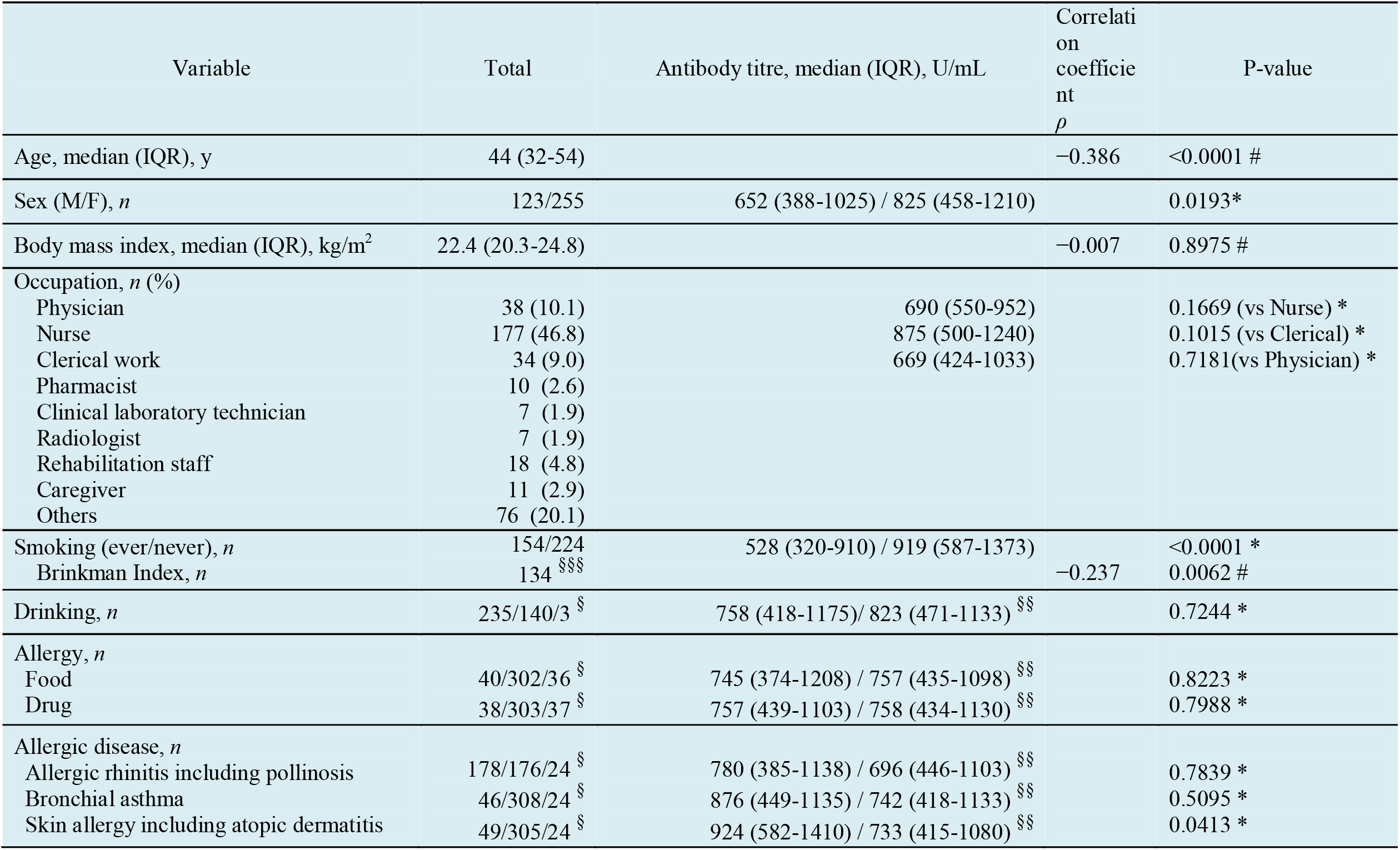
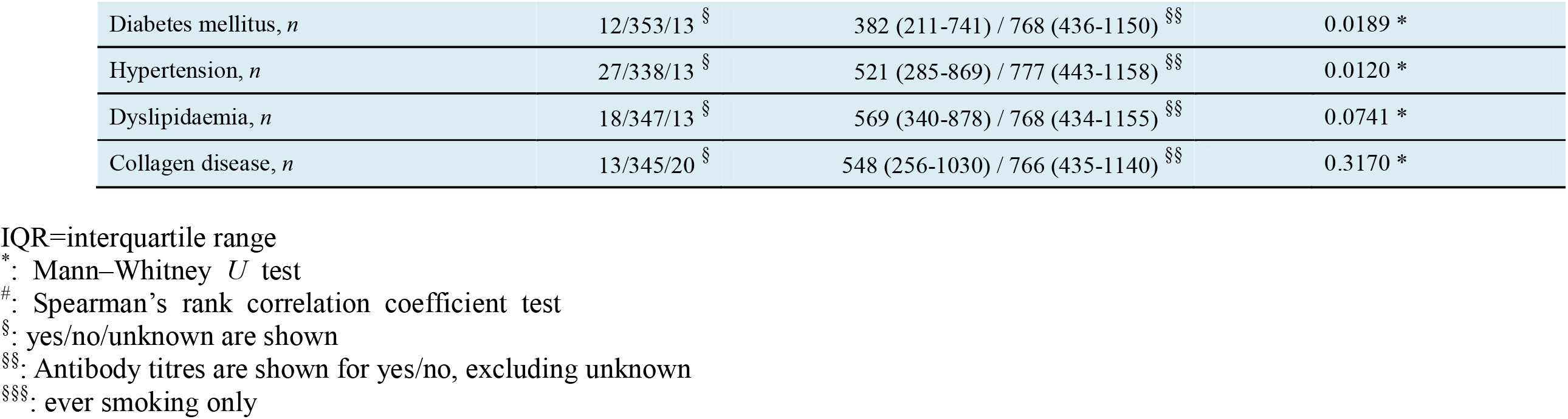
Baseline characteristics of participants (*N*=378)

### Distribution of Ab titres against SARS-CoV-2 spike antigen 3 months after vaccination according to age and sex

Median Ab titre (IQR) against the SARS-CoV-2 spike antigen was 764 (423-1140) U/mL (**Table 1**). Older participants were found to have significantly lower SARS-CoV-2 Ab titres (correlation coefficient ρ=−0.386, p<0.0001) (**Figure 1**). The Ab titres in both men and women tended to decrease with increasing from their 20s to 70s (**Figure 2**). Median (IQR) Ab titres of men in their 20s, 30s, 40s, 50s and 60s-70s were 942 (675-1390), 821 (484-1115), 710 (393-938), 449 (289-861) and 490 (297-571) U/mL, respectively. Median (IQR) Ab titres of women in their 20s, 30s, 40s, 50s and 60s-70s were 1095 (741-1613), 991 (613-1410), 827 (501-1150), 685 (377-1035) and 519 (285-761) U/mL, respectively. Men in each age group tended to have lower median Ab titres than women in the same age group, although the differences were not significant (**Figure 2**).

**Figure 1.**
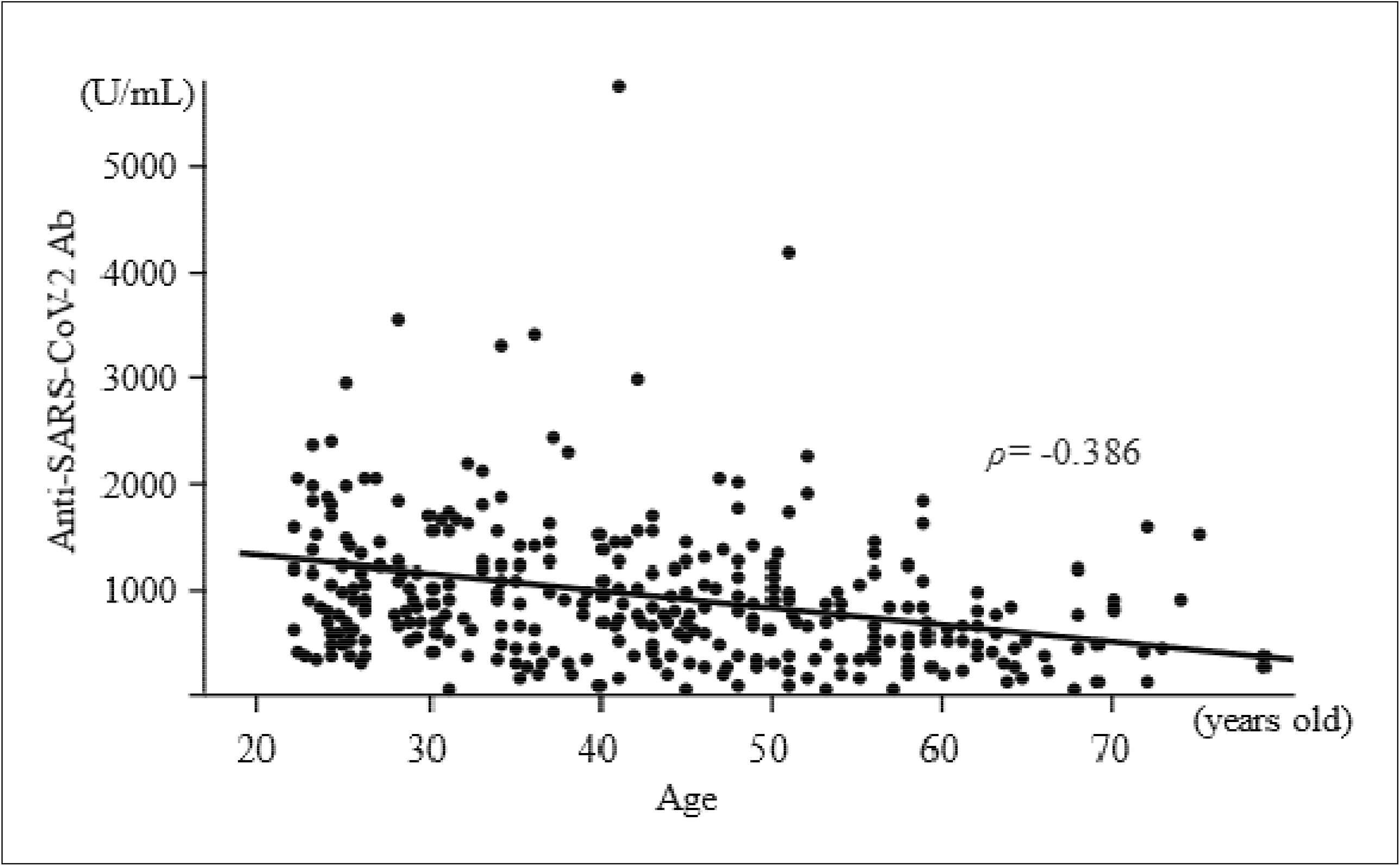
Scatter plot of the distribution of antibody titres according to age. Median antibody titre (IQR) against the SARS-CoV-2 spike antigen was 764 (423-1140) U/mL. Older participants had significantly lower SARS-CoV-2 antibody titres (correlation coefficient ρ=−0.386).

**Figure 2.**
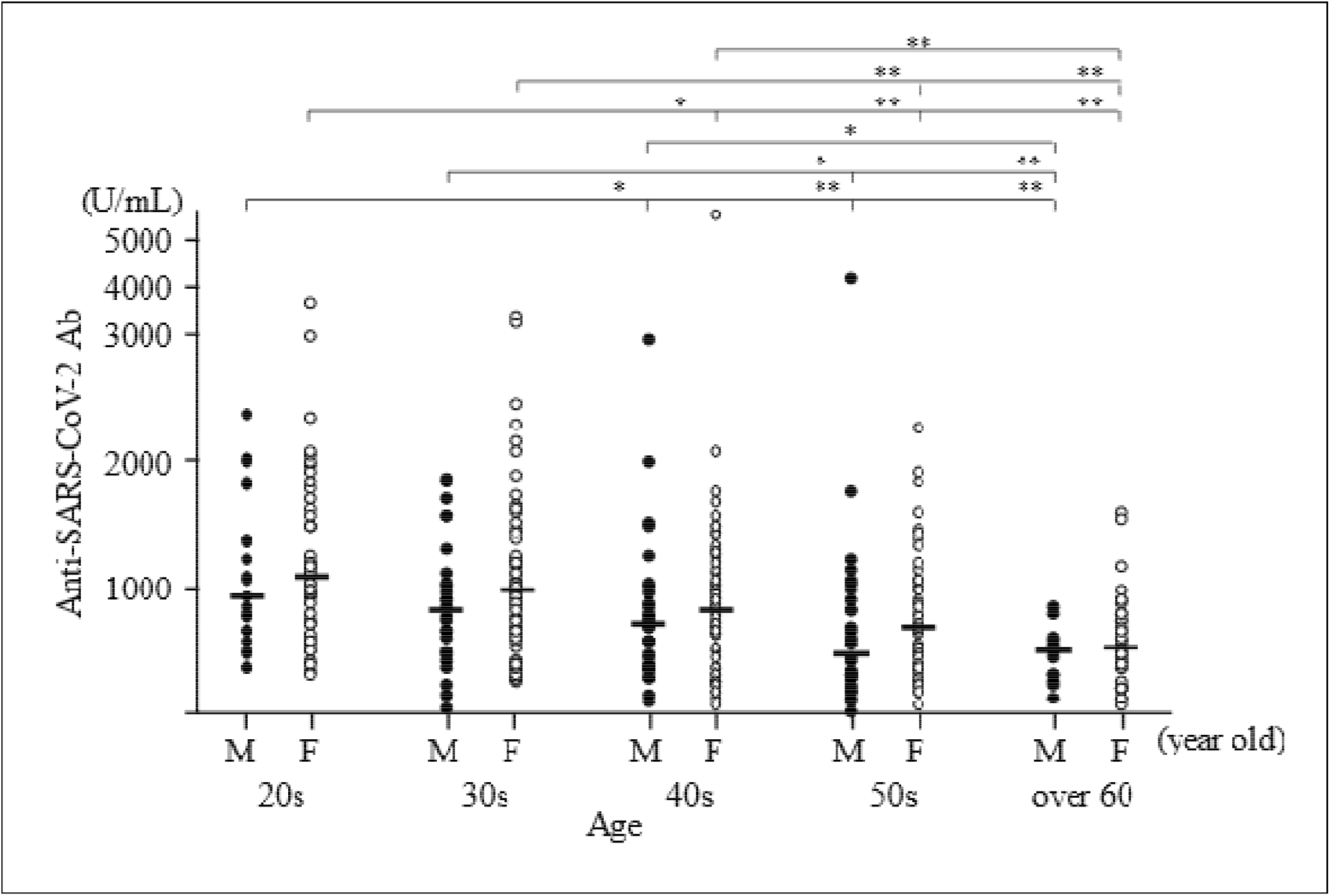
Distribution of antibody titres according to age group and sex. Antibody titres in both men and women tended to decrease as participants’ age increased from their 20s to 70s. Median (IQR) antibody titres of men in their 20s, 30s, 40s, 50s and 60s-70s were 942, 821, 710, 449 and 490 U/mL, respectively. Median (IQR) antibody titres of women in their 20s, 30s, 40s, 50s and 60s-70s were 1095, 991, 827, 685 and 519 U/mL, respectively. Men in each age group tended to have lower median antibody titres than women in the same age group. *p < 0.05, **p < 0.01.

### Relationship between Ab titres against SARS-CoV-2 spike antigen 3 months after vaccination and risk factors

We first performed univariate analyses to identify factors associated with serum Ab titres against the SARS-CoV-2 spike protein. The factors significantly associated with a lower Ab titre were older age, male sex, smoking, skin allergy (including atopic dermatitis), diabetes mellitus and hypertension (**Table 1**).

We also analysed the risk factors for lower Ab titres after adjustment for age, because the prevalence of the factors may differ according to age, such as hypertension. In the age-adjusted analysis, individual Ab titres were recalculated by means of subtraction of the median Ab titres according to corresponding age groups. Median Ab titres of participants in their 20s, 30s, 40s, 50s and 60s-70s were 1050, 931, 779, 619 and 491 U/mL, respectively. For example, an age-adjusted Ab titre in 20s was calculated as follows; ‘an individual Ab titre - 1050’. After age adjustment, the only factors significantly associated with lower Ab titres were male sex and smoking (**Table 2**). In terms of smoking, age-adjusted median Ab titres (IQR) were -174 (−378 to 145) and 90 (−174 to 512) in ever smokers and never smokers, respectively.

**Table 2.** Age-adjusted data of median antibody titres (*N*=378)

| Variable | Total | Antibody titres, median (IQR) <sup>§</sup> | Correlation coefficient $\rho$ | P-value |
| --- | --- | --- | --- | --- |
| Male/female | 123/255 | −69 (−373 to 216) / 55 (−266 to 426) |  | 0.0170 * |
| Occupation, *n* (%) |  |  |  |  |
| Physician | 38 (10.1) | 31 (−263 to 144) |  | 0.6069 (vs Nurse) * |
| Nurse | 177 (46.8) | 46 (−335 to 440) |  | 0.5018 (vs Clerical) * |
| Clerical work | 34 (9.0) | −91 (−309 to 301) |  | 0.7479 (vs Physician) * |
| Smoking (ever/never), *n* | 154/224 | −174 (−378 to 145) / 90 (−174 to 512) |  | <0.0001 * |
| Brinkman Index | 134 <sup>§§§</sup> |  | −0.003 | 0.97010 # |
| Drinking, *n* | 235/140/3 <sup>§</sup> | −18 (−329 to 371) / 32 (−267 to 318) <sup>§§</sup> |  | 0.4955 * |
| Allergy, *n* |  |  |  |  |
| Food | 40/302/36 <sup>§</sup> | 36 (−393 to 357) / −7 (−296 to 319) <sup>§§</sup> |  | 0.7336 * |
| Drug | 38/303/37 <sup>§</sup> | 114 (−165 to 317) / −15 (−309 to 320) <sup>§§</sup> |  | 0.3018 * |
| Allergic disease, *n* |  |  |  |  |
| Allergic rhinitis including pollinosis | 178/176/24 <sup>§</sup> | 10 (−334 to 331) / −19 (−289 to 320) <sup>§§</sup> |  | 0.8541 * |
| Bronchial asthma | 46/308/24 <sup>§</sup> | 93 (−324 to 329) / −23 (−303 to 321) <sup>§§</sup> |  | 0.6102 * |
| Skin allergy including atopic dermatitis | 49/305/24 <sup>§</sup> | 129 (−257 to 522) / −28 (−308 to 299) <sup>§§</sup> |  | 0.0826 * |
| Diabetes mellitus, *n* | 12/353/13 <sup>§</sup> | −169 (−315 to 82) / 0 (−300 to 342) <sup>§§</sup> |  | 0.3105 * |
| Hypertension, *n* | 27/338/13 <sup>§</sup> | −88 (−361 to 284) / 0 (−286 to 338) <sup>§§</sup> |  | 0.3771 * |
| Dyslipidaemia, *n* | 18/347/13 <sup>§</sup> | 24 (−280 to 114) / −3 (−300 to 340) <sup>§§</sup> |  | 0.7484 * |
| Collagen disease, *n* | 13/345/20 <sup>§</sup> | −90 (−516 to 252) / 0 (−299 to 334) <sup>§§</sup> |  | 0.3085 * |

IQR=interquartile range
\*: Mann–Whitney *U* test

To analyse the observed sex difference, we divided the participants into ever smokers and never smokers. In both the male and female groups, age-adjusted median Ab titres were significantly lower in ever smokers than in never smokers; age-adjusted median Ab titres (IQR) in men were -246 (−398 to 65) and 49 (−186 to 621) in ever smokers and never smokers, respectively, while those in women were -140 (−304 to 217) and 95 (−151 to 503) in ever smokers and never smokers, respectively. However, in both the ever smoker and never smoker groups, no significant sex differences in age-adjusted median Ab titres were observed (**Table 3**). Given that the smoking rates in the male and female groups were 61.0% and 31.0%, respectively, these results suggest that the sex difference in Ab titres strongly reflects sex differences in smoking, rather than biological sex differences. In addition, Ab titres were significantly lower in current smokers than in ex-smokers (**Table 4**). These results suggest that smoking cessation will reduce the risk of a lower Ab titre.

**Table 3.** Age-adjusted median antibody titres in ever smokers vs never smokers.

|  | Male,<br>Antibody, median (IQR) | Female,<br>Antibody, median (IQR) | P-value between sexes |
| --- | --- | --- | --- |
| Ever smokers (men, 75; women, 79) | −246 (−398 to 65) | −140 (−304 to 217) | 0.1175 * |
| Never smokers (men, 48; women, 176) | 49 (−186 to 621) | 95 (−151 to 503) | 0.9970 * |
| P-value between ever and never smokers | 0.0007 * | 0.0023 * |  |

IQR=interquartile range
\*: Mann–Whitney *U* test

**Table 4.**
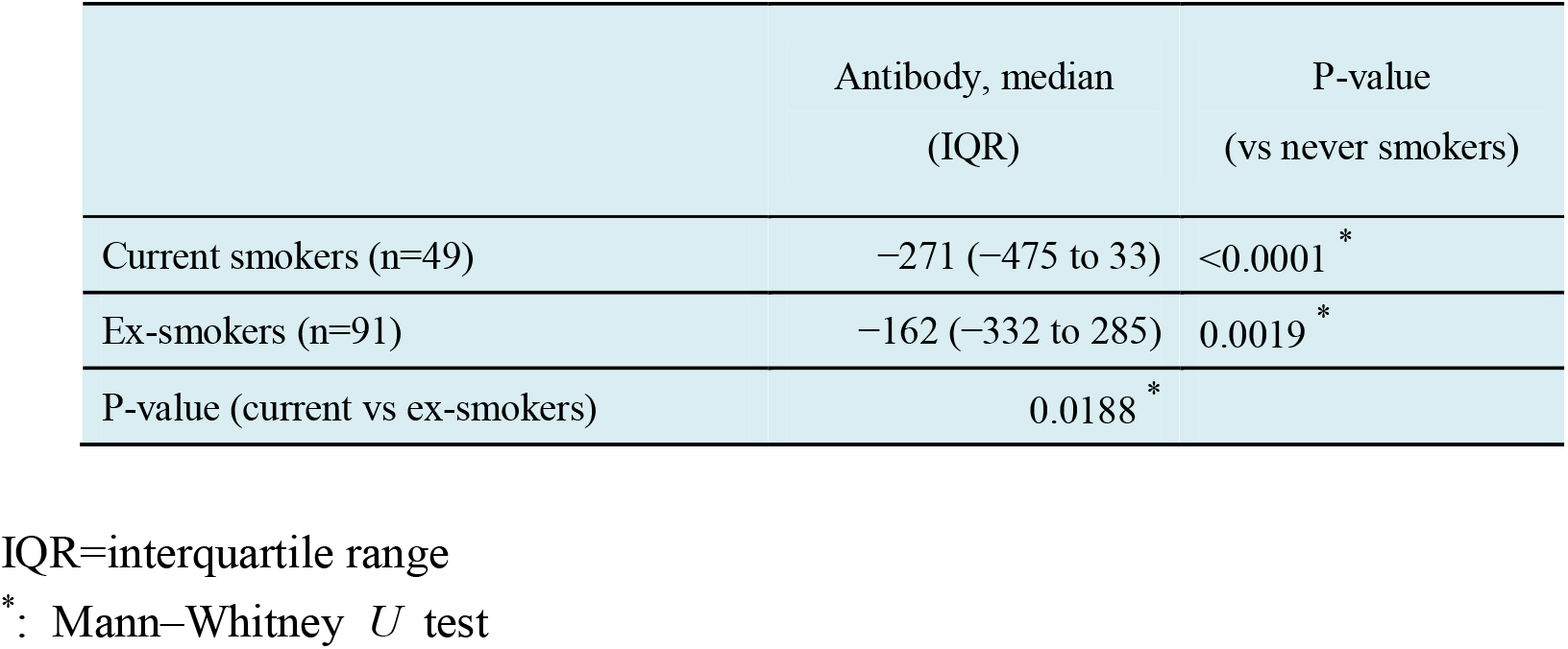
Age-adjusted median antibody titres in ever smokers: current smokers vs ex-smokers.

## DISCUSSION

To our knowledge, this is the first study to report real-world Ab titres against the SARS-CoV-2 spike antigen at 3 months after vaccination and to identify the factors associated with these Ab titres across a comprehensive range of clinical and lifestyle characteristics in Japan. Three important findings were obtained. First, median age (IQR) of the participants was 44 (32-54) years, median Ab titre (IQR) against the SARS-CoV-2 spike antigen was 764 (423-1140) U/mL, and older participants had significantly lower Ab titres, with median (IQR) Ab titres of 942 (675-1390) and 1095 (741-1613) U/mL in men and women in their 20s, respectively, but 490 (297-571) and 519 (285-761) U/mL in men and women in their 60s-70s, respectively. Second, in the age-adjusted analysis, the only risk factors for lower Ab titres were male sex and smoking. However, the sex difference may have arisen from the sex difference in smoking rate. Third, Ab titres were significantly lower in current smokers than in ex-smokers. Smoking cessation can thus be expected to reduce the risk of lower Ab titres.

Because the efficacy of COVID-19 mRNA vaccines in clinical trials and their effectiveness in the community depend on the proportions of SARS-CoV-2 variants spreading in a given area, immunogenicity has attracted increasing attention as an individual index for the efficacy of these vaccines. Neutralising Abs are correlated with protection against SARS-CoV-2 [5], but only a few studies have investigated real-world Ab titres following vaccination with BNT162b2, focusing instead on Ab status shortly after vaccination. Those studies demonstrated that lower Ab titres may be caused by older age [10], male sex [10], obesity [14, 15], smoking habit [15], drinking habit [16], hypertension [15], cancer [16, 17], use of immunosuppressive drugs [10] and a longer time elapsed after vaccine inoculation [10, 15]. Medium-term data of serological Ab titres in response to the BNT162b2 vaccine are urgently needed, as are the clinical and lifestyle factors predicting these titres.

Concerning our first main finding, the median titre in individuals shortly after a full vaccination schedule of this vaccine in Japan was reported to be 2060 U/mL (IQR, 1250-2650) [10], which is similar to the median titre reported from Italy (1975 U/ml; IQR, 895-3455) [20]. However, anti-SARS-CoV-2 Ab titres following vaccination cannot currently predict the likelihood of developing COVID-19. Our Ab titres 3 months after the second inoculation ranged from 3 to 5790 U/mL and the median Ab titre (IQR) was 764 (423-1140) U/mL, which was much lower than the above-mentioned value obtained shortly after the second inoculation, reflecting the reported association between longer time elapsed since the second inoculation and lower Ab titres. Moreover, older participants had significantly lower SARS-CoV-2 Ab titres. Indeed, in our preliminary study [18] with 6 participants in their 50s and 60s, the average peak titre of 2031.7±692.0 U/mL had markedly decreased to 513.3±261.7 U/mL by 15 weeks after the second inoculation. Two of the 6 subjects had Ab titres of 220.0 U/mL, which is a similar level to that observed immediately before the second inoculation, suggesting that vaccination with COVID-19 mRNA vaccine may be needed every 6 months for people over 50 years old in Japan [18]. Although sufficient clinical efficacy of the BNT162b2 vaccine against most of the conventional SARS-CoV-2 variants was observed even after 6 months without Ab titre data [21], the clinical efficacy of a single inoculation with this vaccine against the SARS-CoV-2 Delta variant has been reported to be only 33.5% [22].

Concerning our second main finding, various reports have demonstrated that older age [10-12], male sex [10], obesity [14, 15], smoking habit [15], drinking habit [10], hypertension [15] and cancer [16, 17] may reduce Ab responses to COVID-19 mRNA vaccination shortly after the second inoculation, in addition to a longer time elapsed after vaccine inoculation [10, 15] and use of immunosuppressive drugs [10]. Our participants receiving immunosuppressive drugs also seemed to have low Ab titres, including a man in his 30s taking infliximab [23] with a titre of 9.0 U/mL, a woman in her 40s taking etanercept with 258 U/mL and a woman in her 30s taking etanercept with 415 U/mL. In our study, smoking was the most impactful factor, and the sex difference in Ab titres was at least partly due to smoking. To clarify the effects of smoking, we performed additional analyses. However, the Brinkman Index and the number of cigarettes per day did not influence the Ab titres. Thus, smoking itself is a risk factor for low Ab titres, rather than the duration of smoking or number of cigarettes per day. Moreover, smoking cessation can be expected to effectively increase Ab titres because they were significantly lower in current smokers than in ex-smokers. On the other hand, another study reported that alcohol was the most impactful lifestyle factor, not smoking [10]. Although the authors of that study do not show age-adjusted data, their conclusions were different from ours. Their results were based on analysis of samples collected 2-5 weeks after the second dose of the vaccine. In contrast, our samples were obtained 3 months after the second dose. One hypothesis is that alcohol may affect Ab production, whereas smoking may lead to decreases in the level of Abs after they are produced. In addition, we believe that an age-adjusted analysis is critical. To clarify the reason for the difference between that study and ours, we need to analyse longitudinal data to observe changes in Ab titres over time, and we are planning such an additional study. Although the mechanisms are not known, previous studies have reported an association between a smoking habit and lower Ab titres against both influenza virus [24] and hepatitis B virus [25] following vaccination.

Some limitations and possible sources of bias in this study include the following. First, the participants were limited in number and were all healthcare workers vaccinated at a single national hospital in Tochigi prefecture, where the COVID-19 pandemic has been well-controlled since SARS-CoV-2 began spreading around the globe. Therefore, the results obtained in this study might not be generalisable on a wide scale, or even within Japan. Second, several data, including body height and weight, were obtained by means of a standardised structured self-reported questionnaire. We cannot deny the possibility that some data may have been affected by recall bias. Third, the cut-off level of the Ab against the SARS-CoV-2 spike antigen needs to be determined for the different variants of SARS-CoV-2, but this information is currently unavailable. Further studies are needed to confirm our observations.

In conclusion, we have reported for the first time real-world Ab titres against the SARS-CoV-2 spike antigen at 3 months after the second dose of the BNT162b2 vaccine, which were much lower than those measured shortly after the second inoculation. We demonstrated that the most important factors associated with low Ab titres were age and smoking habit. In particular, current smoking status causes lower Ab titres, and smoking cessation before vaccination may improve the individual effectiveness of the BNT162b2 vaccine. To establish a more personalised approach to vaccination involving earlier boosters, different schedules or different types of vaccines, further studies are necessary regarding the associations between Ab titres and the comprehensive medical histories of individuals.

## Data Availability

All data referred to in the manuscript are available.

## ACKNOWLEDGEMENTS

The authors thank all staff in National Hospital Organization Utsunomiya National Hospital for supporting sample collection. We would like to thank Ms. Miwa Okada, Ms. Junko Shibayama, Ms. Yuko Tajima, Ms. Mami Ochiai, Ms. Midori Takahashi, Ms. Hiroko Ueno, Ms. Natsuka Suzuki, and Ms. Yoshino Iwaya for supporting data analysis.

## DISCLOSURE STATEMENT

The authors declare no conflict of interest.

## Notes

### Competing Interest Statement

The authors have declared no competing interest.

### Funding Statement

No fundings.

### Author Declarations

This study was approved by the Ethics Committee of National Hospital Organization Utsunomiya National Hospital (No. 03-01, April 19, 2021).

